# Comparative Evaluation of Time Series Forecasting Approaches for Facility-Level Antibiotic Resistance Outcomes in the Veterans Health Administration

**DOI:** 10.1101/2025.04.17.25325964

**Authors:** Haojia Li, Matthew H. Samore, Yue Zhang

**Affiliations:** Informatics, Decision Enhancement, and Analytic Sciences Center (IDEAS), VA Salt Lake City Healthcare System, Salt Lake City, Utah, USA; Division of Epidemiology, Department of Internal Medicine, University of Utah, Salt Lake City, Utah, USA; Division of Biostatistics, Department of Population Health Sciences, University of Utah School of Medicine, Salt Lake City, Utah, USA

## Abstract

Antibiotic resistance is a critical public health threat, particularly in hospital settings where vulnerable populations face heightened risks of infection and adverse outcomes. Forecasting resistance trends at the facility level is essential for guiding local antibiotic stewardship, informing infection control strategies, and improving patient safety. In this study, we conducted a comparative evaluation of time series forecasting methods to predict facility-level antibiotic resistance trends in hospital-onset infections within the Veterans Health Administration system. Using data from January 2007 to March 2022 across 30 high-volume facilities, representing approximately half of all admissions and patient days, we assessed the performance of autoregressive integrated moving average (ARIMA), vector autoregression (VAR), and long short-term memory (LSTM) models. Our results showed that LSTM models consistently outperformed ARIMA and VAR, with accuracy notably enhanced by incorporating antibiotic use covariates. These findings underscore the value of integrating antimicrobial utilization data and deep learning approaches to improve resistance forecasting and support targeted stewardship efforts.

## 1 Introduction

Antibiotic resistance is a growing public health crisis, particularly in healthcare settings where vulnerable populations are at heightened risk for infection and adverse outcomes ^1^. Hospital-onset infections (HOIs), often involving multidrug-resistant organisms (MDROs), represent a significant burden on patients and healthcare systems, increasing morbidity, mortality, and healthcare costs ^2,3^. Monitoring and forecasting antibiotic resistance trends at the facility level is critical for guiding local antibiotic stewardship practices, informing infection control strategies, and improving patient safety outcomes ^4,5^.

The Veterans Health Administration (VHA), the largest integrated healthcare system in the United States, provides a unique opportunity to examine facility-level variation in antimicrobial resistance across a diverse national network of hospitals ^6,7^. Accurate, forward-looking predictions of antibiotic resistance rates in VHA facilities can support tailored interventions that reflect local epidemiology and prescribing patterns. Despite the recognized importance of local surveillance, most existing models for resistance forecasting operate at national or regional levels, potentially obscuring the granular trends necessary for effective facility-specific decision-making ^8^.

Time series forecasting models, including statistical approaches like autoregressive integrated moving average (ARIMA), as well as machine learning and deep learning methods, have shown utility in predicting healthcare outcomes and antimicrobial resistance ^9–11^. In recent years, deep learning models such as Long Short-Term Memory (LSTM) networks have demonstrated superior performance in capturing temporal dependencies and non-linear patterns in health data ^12,13^. However, their comparative performance for facility-level prediction of drug-specific antibiotic resistance, particularly in the context of hospital-onset infections, remains underexplored. Furthermore, antibiotic utilization is a well-established driver of resistance, yet few studies have evaluated how drug use profile impact the accuracy of resistance forecasts ^14,15^.

In this study, we conducted a comparative evaluation of time series forecasting approaches, including ARIMA, Vector Autoregression (VAR) and LSTM, for predicting facility-level antibiotic resistance trends in HOIs within the VHA. Our findings aim to support data-driven antimicrobial stewardship strategies tailored to individual healthcare facilities.

## 2 Methods

### 2.1 Study Design

We conducted a retrospective observational study using data on inpatient antimicrobial susceptibility test (AST) results and antibiotic usage in the national electronic health records (EHR) of the VHA. The study period spanned from January 1, 2007, to March 31, 2022, covering a total of 183 months. We prioritized VHA facilities with a high total number of inpatient admissions during this period to ensure sufficient data volume for robust time series modeling. We aggregated monthly AST results into multiple facility-level antibiotic resistance rate time series. Our objective was to compare various time series forecasting models for monthly facility-level antibiotic resistance rates, with the goal of informing data-driven antimicrobial stewardship and supporting local resistance surveillance efforts.

### 2.2 Data

We collected data on nine clinically relevant pathogens: *Acinetobacter sp*., *Enterobacter cloacae (E. cloacae)*, *Escherichia coli (E. coli)*, *Enterococcus faecalis (E. faecalis)*, *Enterococcus faecium (E. faecium)*, *Klebsiella pneumoniae (K. pneumoniae)*, *Pseudomonas aeruginosa (P. aeruginosa)*, *Serratia marcescens (S. marcescens)*, and *Staphylococcus aureus (S. aureus)*. AST results were categorized as susceptible, intermediate, or resistant. We restricted analyses to incident isolates, which were defined as the first isolate per patient–pathogen pair within a 30-day period, to reduce bias from repeated sampling and ensure comparability across facilities. To distinguish hospital-acquired resistance from resistance present at the time of admission, we categorized infections as either HOIs or community-onset infections (COIs), based on the timing of specimen collection relative to admission. We defined resistance as the presence of either a resistant or intermediate AST result and calculated resistance rates as the number of resistant isolates per 1,000 patient-days, aggregated monthly by pathogen and facility.

In this study, we focused on four facility-level monthly antibiotic resistance outcomes for HOIs, selected based on clinical importance and data availability: methicillin-resistant *S. aureus* (MRSA), fluoroquinolone-resistant *S. aureus*, third- or fourth-generation cephalosporin-resistant *E. coli*, and fluoroquinolone-resistant *E. coli*. We combined third- and fourth-generation cephalosporins into a single antibiotic category due to their similar spectrum of activity and common use in empiric treatment of Gram-negative infections. To define resistance for this class, we considered an isolate resistant if it was classified as resistant or intermediate to at least one agent in either generation. This approach minimized overestimation of resistance risk compared to aggregating resistance rates across agents, particularly in cases where both cephalosporins were tested in the same isolate.

To improve forecasting accuracy and incorporate potential signals relevant to resistance dynamics, we considered five groups of monthly facility-level predictors that could potentially influence resistance dynamics:

1. Antibiotic use rates Monthly inpatient antibiotics use, measured in per 1,000 patient-days. This group included the antibiotic classes corresponding to the resistance outcome, as well as related classes that may exert indirect selective pressure.
2. HOI incidence for the outcome pathogen The incidence rate of HOIs caused by the same pathogen as the outcome, calculated as the number of HOIs per 1,000 admissions.
3. Resistant or susceptible COI incidence for the outcome pathogen and antibiotic class The incidence rate of COIs involving the same pathogen–antibiotic-class pair as the outcome, including both resistant and susceptible phenotypes.
4. Resistant HOI incidence for the outcome pathogen and related antibiotic classes The incidence rate of resistant HOIs involving the same pathogen and related antibiotic classes, expressed per 1,000 patient-days.
5. Resistant HOI incidence for the outcome antibiotic class and related pathogens The incidence rate of resistant HOIs involving related pathogens and the same antibiotic class. This group was designed to capture cross-pathogen resistance pressure and was also expressed per 1,000 patient-days.

Table 1 summarizes the predictors and indicates their relevance for each specific resistance outcome based on clinical relevance. For modeling purposes, we constructed lagged versions of each predictor using lag orders 1 through 12 to capture potential delayed effects on resistance outcomes.

**Table 1:**
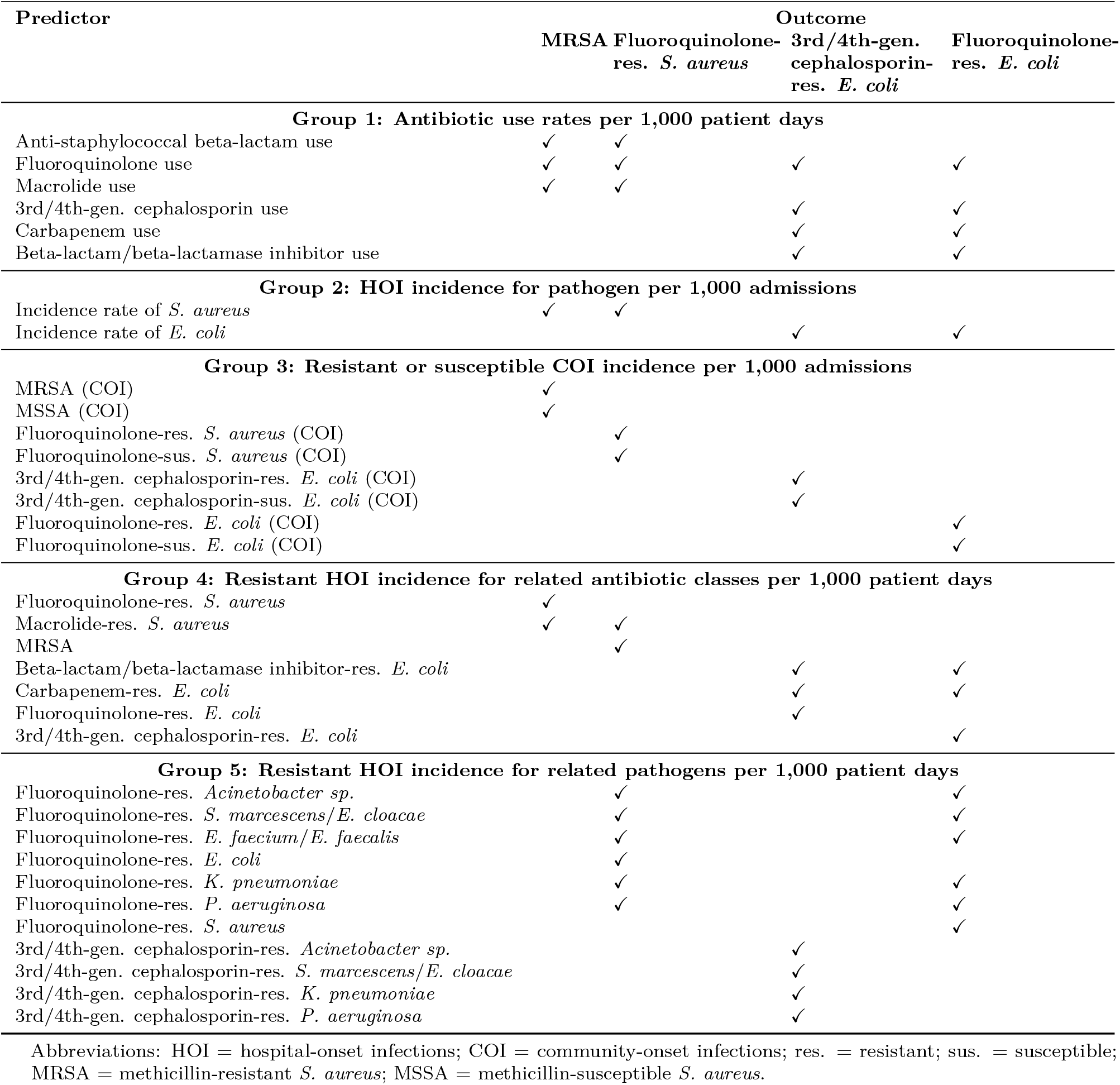
Facility-level predictors grouped by relevance to resistance outcomes.

Given the granularity of the data, which were aggregated at the facility–month level, many time series contained months with zero observed isolates, representing periods without reported cases rather than true missingness. While we treated missing values as zeros, we conducted visual inspection of all outcome and predictor time series using line charts to identify irregular patterns such as discontinuities, implausible fluctuations, or prolonged flat segments. These checks helped detect potential data quality issues that may not be captured by summary statistics alone.

To ensure meaningful modeling and adequate data availability, we calculated the proportion of missing values for each variable, and excluded outcome time series in which more than 70% of facility-month values were zero. For predictors, we applied a more lenient threshold of 90%. These thresholds were selected to balance data retention with the need for reliable inputs in downstream modeling.

### 2.3 Time Series Decomposition and Data Preprocessing

We used the seasonal-trend decomposition using LOESS (STL) ^16^ to quantify the presence and strength of trend and seasonal components in each time series. We performed an additive decomposition, in which the observed data can be expressed as:

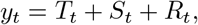

where *y*_*t*_ is the observed data, *T*_*t*_ is the trend-cycle component, *S*_*t*_ is the seasonal component, and *R*_*t*_ is the residual component, all at time *t*.

To assess the contribution of trend and seasonality to the overall variability in each series, we computed the strength of trend (*F*_*T*_) and strength of seasonality (*F*_*S*_) using the following metrics^17^:

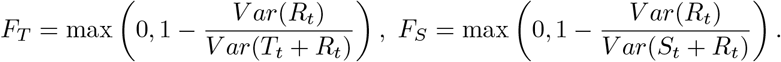

We interpreted the strength of trend and seasonality in 0-0.3 as weak, 0.3-0.6 as moderate, and 0.6-1 as strong, and considered seasonality relevant for further modeling only when its strength was classified as strong.

To ensure that the time series met the assumptions required for subsequent modeling, we conducted stationarity diagnostics using both the Augmented Dickey–Fuller (ADF) test and the Kwiatkowski–Phillips–Schmidt–Shin (KPSS) ^18^ test. The following sequential procedure was applied to each series:

1. Standardize the data by subtracting the mean and dividing by the standard deviation.
2. Apply the ADF and KPSS tests to the standardized data.
3. If either test indicated non-stationarity, apply first-order differencing and repeat step 2.
4. If non-stationarity persisted, apply STL-based detrending and re-evaluate stationarity.

These steps were designed to produce stationary series appropriate for statistical time series modeling, while preserving interpretable trend and seasonal structures when relevant.

### 2.4 Model Specification and Parameter Tuning

We used three types of time series models: ARIMA, VAR and LSTM. The ARIMA model comprises the autoregressive (AR), differencing, and moving average (MA) components, corresponding to the parameters *p, d*, and *q*, respectively. VAR generalizes the univariate autoregressive model to a multivariate framework, using a single lag order *p* to model interdependencies among multiple time series. We used ARIMA to capture linear autocorrelated structures within individual time series, and VAR to account for potential cross-correlations among the outcomes. We conducted parameter tuning for ARIMA and VAR models, as described in this section later. LSTM, a type of recurrent neural network (RNN), is well-suited for modeling complex temporal dependencies, including nonlinear dynamics and long-range interactions ^19^. Our LSTM architecture adopted a sequence-to-one structure, mapping a fixed-length input sequence to a single-step prediction at each iteration. Across all models, we implemented a one-step-ahead forecasting framework, wherein predictions at each time point were generated based solely on information available up to the previous time step.

We conducted parameter tuning for ARIMA and VAR. For each stationary time series, we plotted autocorrelation function (ACF) and partial autocorrelation function (PACF) up to lag of 12 months to determine the range of parameters *q* and *p* in parameter tuning for ARIMA, respectively. The minimum of both *p* and *q* was 0. The maximum of *p* was determined as the largest lag where the PACF was statistically significant at the 95% confidence level, plus 1 as a buffer, with a lower bound of 3 and an upper bound of 12. The maximum of *q* was chosen using the same approach based on ACF. The possible values of *d* were chosen from 0 and 1. We performed a grid search over the possible values of *p, d*, and *q* to find the optimal parameters. For VAR, we uniformly included the lag order from 1 to 12 in parameter tuning.

An expanding window structure was used to conduct the parameter tuning (Figure 1), where the training set started with size of 12 months, and progressively expanded by 1 month each time, while the validation set remained fixed at 3 months following the training set. The choice of a 12-month initial training window allows the model to capture a full seasonal cycle and provides sufficient historical context for stable learning. Expanding the training set by 1 month at a time reflects a realistic model updating strategy and enables fine-grained performance evaluation over time. A 3-month validation window balances short-term forecast relevance with sufficient data for robust validation. In each window, we fitted the model on the training set, and evaluated the model performance on the validation set using the root mean squared error (RMSE). The optimal parameters were chosen based on the smallest average RMSE across all windows.

**Figure 1:**
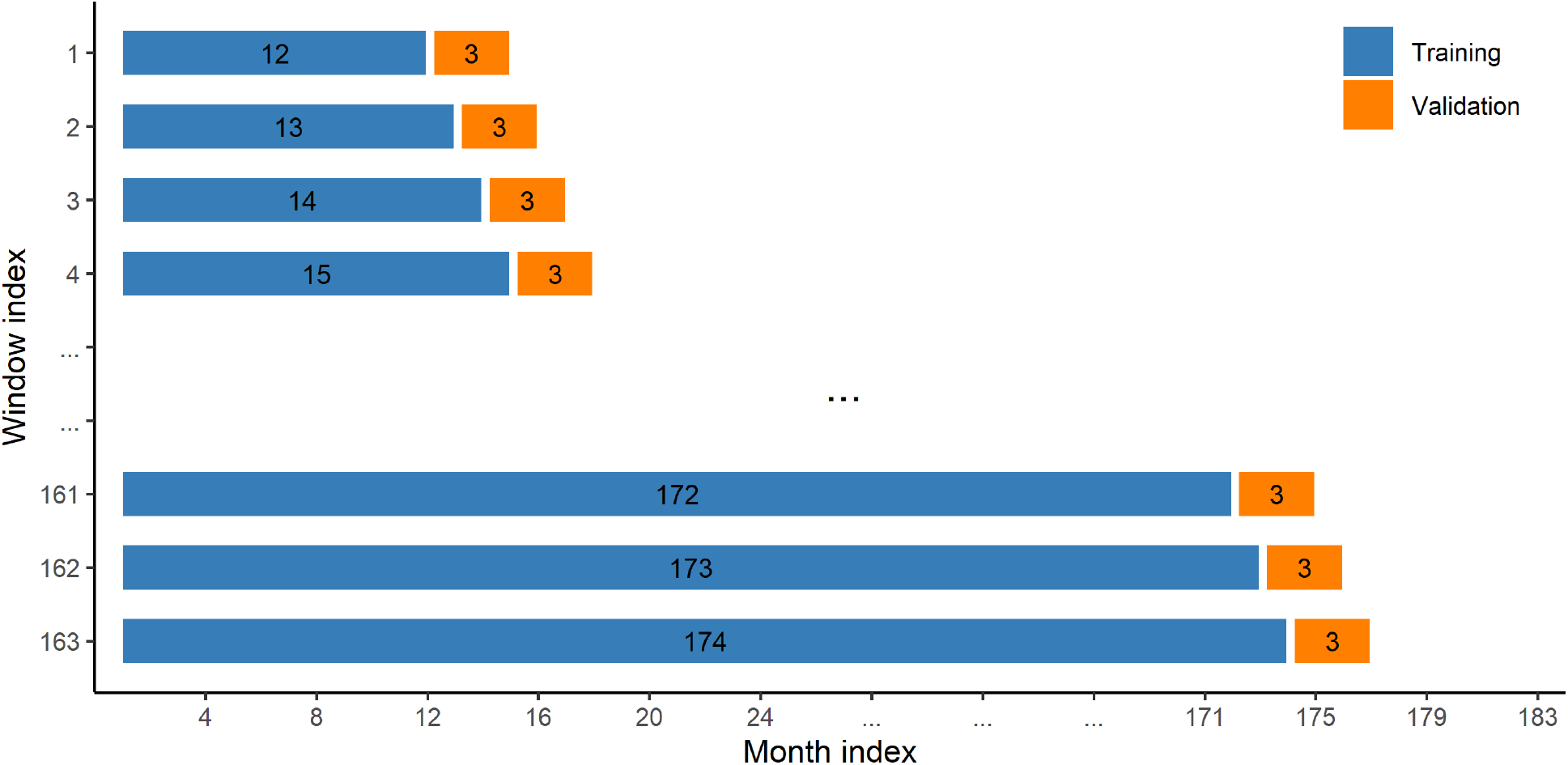
Data partitioning strategy for model tuning, calibration, and evaluation. Expanding window framework used for parameter tuning. Each row represents one window, with bar segments showing training and validation sets. Numbers in each segment indicate the number of months included.

We selected LSTM hyperparameters based on prior work and initial experimentation, rather than performing a full grid search. We set the input sequence length to 12 time steps and configured the model to predict one time step ahead at each iteration. The RNN module includes two stacked LSTM layers, each with 32 hidden units, a configuration chosen to balance model capacity and generalizability. To mitigate overfitting, we applied a dropout rate of 0.2 between layers. We trained the model for up to 300 epochs using a batch size of 16 and a learning rate of 0.001, which yielded stable convergence across expanding window evaluations. To prevent overtraining, we implemented early stopping, halting training if the training loss failed to improve by more than 0.001 over 10 consecutive epochs. We monitored training loss, rather than validation loss, as the short and shifting validation windows introduced variability that could lead to premature stopping.

### 2.5 Final Model Evaluation and Comparison

To compare the predictive performance across different models, we applied the final forecasting models from each approach to predict outcomes from month 25 to 183 and computed the RMSE between the predicted and observed values. As we applied one-step-ahead prediction, the RMSE remained interpretable and comparable across outcomes, regardless of whether first-order differencing was applied during data preprocessing based on each model’s assumptions and configuration. We kept RMSE on the standardized scale to facilitate consistent comparison across time series with different units and magnitudes. We evaluated both outcome-only and predictor-enhanced versions of ARIMA, VAR, and LSTM models. While outcome-only models used only lagged values of the outcome time series, predictor-enhanced models incorporated predictors with lag orders from 1 to 12. To manage the number of predictors and maintain model identifiability, particularly for ARIMA and VAR, we included only antibiotic use rates as predictors in the predictor-enhanced models used to compare the forecasting performance of ARIMA, VAR, and LSTM. To quantify the contribution of antibiotic use rates in the prediction, we computed RMSE ratios by dividing the RMSE from the antibiotic-use–enhanced model by that of the corresponding outcome-only model. A ratio below 1 indicates improved accuracy, with smaller values reflecting greater benefit from predictor inclusion.

To further assess the contribution of antibiotic use variables, we performed zero-out analysis using LSTM models. We used the LSTM models that included all five predictor groups as the benchmark for comparison. First, we zeroed out one predictor group at a time while retaining all others to evaluate the overall importance of each group. We hypothesized that the antibiotic use rates had the greatest contribution to the prediction of resistance outcomes due to its clinical relevance and direct relationship to selection pressure. We excluded Group 5 predictors from this analysis and retained them in all models. We then conducted a second round of zero-out analysis by removing each individual antibiotic use variable one at a time to identify the most influential agents. For each reduced model, we calculated the RMSE ratio relative to the fully predictor model. Higher ratios indicate greater loss in predictive accuracy, and thus greater importance of the excluded group or predictor. We summarized the results across facilities using box plots of RMSE ratios by predictor group and reported the mean RMSE ratio per group to assess overall impact.

### 2.6 Implementation and Software Tools

We used SQL to extract data from the VHA EHR database and to preprocess the initial datasets, including filtering and combining large tables. The majority of the analysis was conducted in R (version 4.4.1), including data extraction (`DBI`, `odbc`), data processing (`tidyverse`), ARIMA (`forecast`) and VAR modeling (`vars`), and results summarization. Grid search for parameter tuning was parallelized using the `parallel` package. LSTM modeling was implemented in Python (version 3.10.16) using the `darts`^20^ time series library.

## 3 Results

### 3.1 Descriptive Summary and Time Series Patterns

We included 30 VHA facilities in the analysis, among which both the number of admissions and the number of patient days accounted for approximately half of the overall volume observed across all 124 VHA facilities during the study period from January 2007 to March 2022. The majority of the selected facilities were located in urban areas and classified as high complexity levels according to VHA facility designations, reflecting the presence of teaching and clinical programs and volume of high-need patients.

After excluding time series with more than 70% missing values, we retained 104 time series across four antibiotic class–pathogen pairs. Figure 2 illustrates the temporal trend in the average outcome across the selected facilities. MRSA and fluoroquinolone-resistant *S. aureus* exhibited clear decreasing trend, with more pronounced declines during the earlier years of the study period. Though the trends were more gradual, third- or fourth-generation cephalosporin-resistant *E. coli* increased slightly over time, while fluoroquinolone-resistant *E. coli* showed a downward trend.

**Figure 2:**
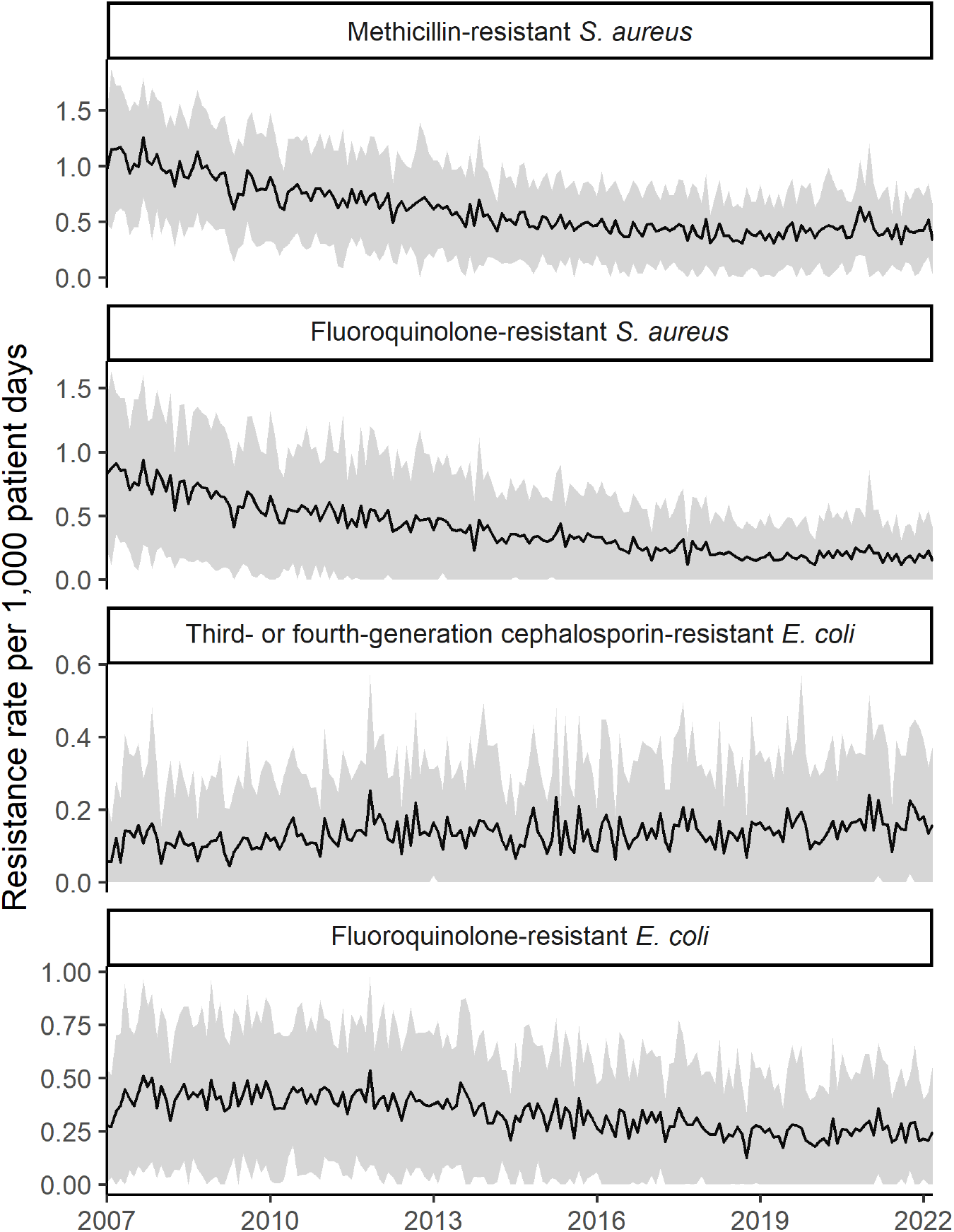
Temporal trend in the average outcome across the selected facilities. The shaded area represents the range of one standard deviation above and below the mean, reflecting the variation in resistance rates. Lower bounds were truncated at zero to avoid negative values.

As shown in Figure 3, the strength of seasonality was weak across most time series, with the majority showing values well below 0.3. The only exception was the macrolide use rate, which exhibited a median strength of seasonality slightly above 0.3. This suggests that seasonality is not a dominant feature in most series and may not require explicit modeling in most cases. The strength of trend showed greater variability. Antibiotic use rate series exhibited the highest overall strength of trend across all groups, indicating strong long-term patterns in prescribing behavior. We observed substantial variation in the strength of trend within each time series group, highlighting the heterogeneity in temporal dynamics across different pathogen, antibiotic class, and pathogen–antibiotic-class combinations. All time series were found to be stationary, as assessed by both ADF and KPSS test, after preprocessing steps up to standardization and, when necessary, first-order differencing.

**Figure 3:**
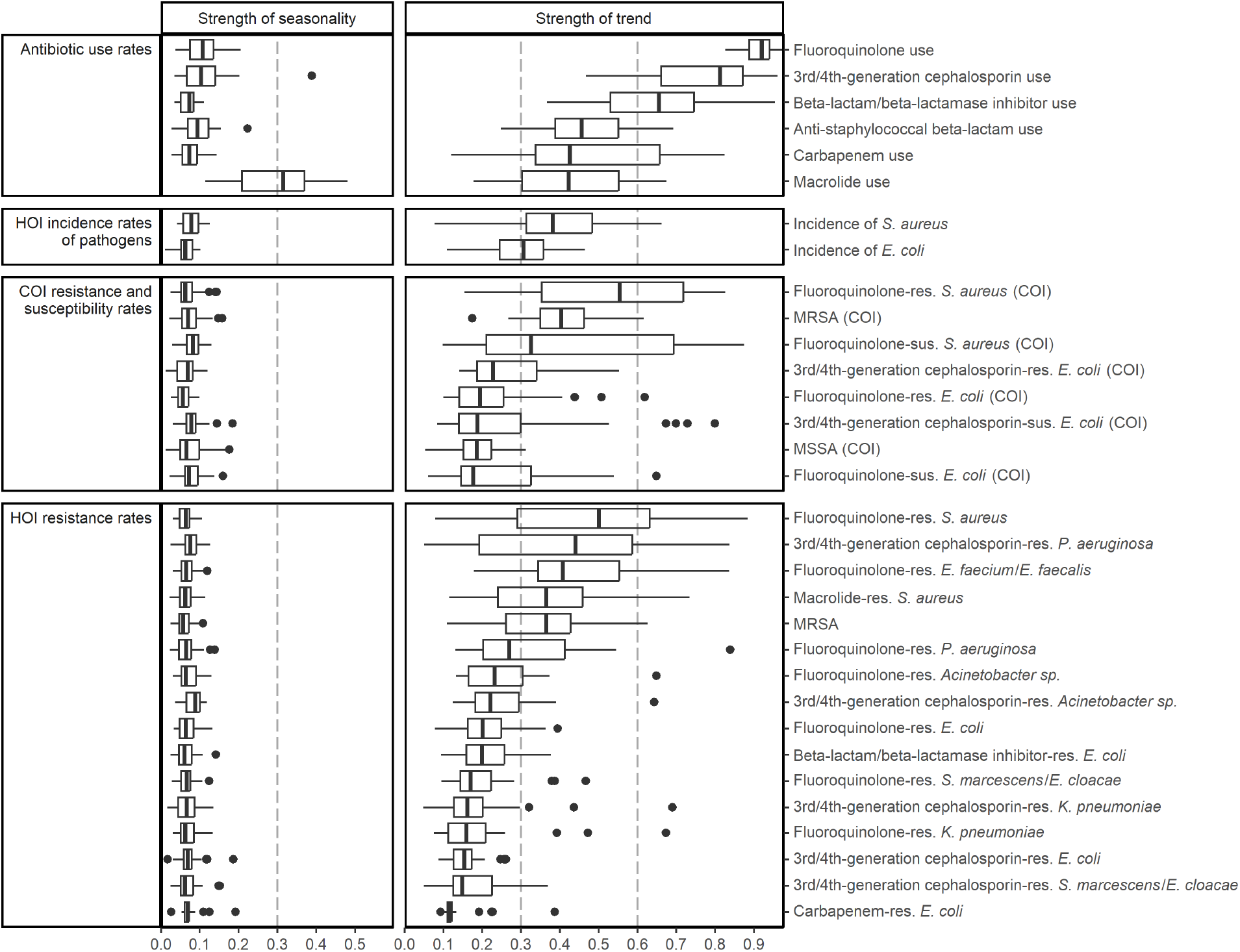
Distribution of strength of seasonality and trend across facilities for different time series in the analysis. Strength values represent the proportion of variation in the time series explained by the seasonal or trend component, based on STL decomposition. Dashed vertical reference lines are drawn at 0.3 and 0.6 to indicate interpretive thresholds for classifying strength values as weak (0–0.3), moderate (0.3–0.6) and strong (0.6–1). Abbreviations: HOI = hospital-onset infections; COI = community-onset infections; res. = resistant; sus. = susceptible; MRSA = methicillin-resistant *S. aureus*; MSSA = methicillin-susceptible *S. aureus*.

ARIMA model tuning across the 104 stationary time series revealed several key patterns. Nine series were best fit by an ARIMA(0,0,0) model, indicating that a constant mean was sufficient to capture the underlying dynamics. Six of these series were for fluoroquinolone-resistant *E. coli*, which aligns with the relatively stable trend observed in the corresponding average outcome series. None of the series selected a pure autoregressive model (ARIMA(*p*, 0, 0) with *p* > 0) as the optimal structure. By contrast, 40 series (38.5%) were best fit by a pure moving average model (ARIMA(0, 0, *q*) with *q >* 0). The majority of models selected *p* = 0 (81 series) or *p* = 1 (19 series), and *q* = 1 (63 series) or *q* = 2 (29 series) were the most frequently selected MA orders. Forty-six series (44.2%) had an optimal model with a differencing term of *d* = 1. We observed variation in the average ARIMA parameters selected during model tuning (Table 2) across the four outcome groups, . In the final tuned models, fluoroquinolone-resistant *S. aureus* had the highest average autoregressive order at 0.70, whereas the other outcomes had average orders between 0.14 and 0.20. In terms of moving average order (*q*), all outcomes except for third- or fourth-generation cephalosporin-resistant *E. coli* had an average tuned value greater than 1, with MRSA exhibiting the highest average at *q* = 1.50. The optimal lag order selected for the VAR model was 1 across all facilities.

**Table 2:**
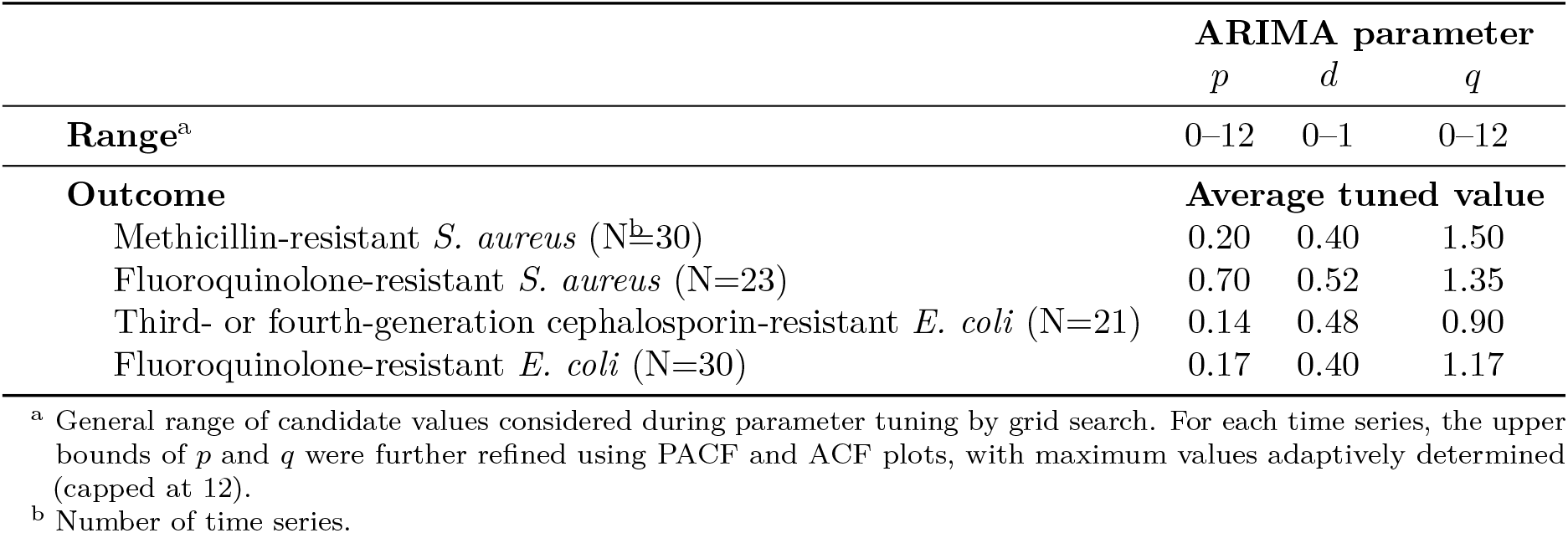
Average ARIMA model parameters by outcome.

### 3.2 Model Performance

With 97 (93.3%) out of 104 outcome time series achieving the lowest RMSE, the LSTM model incorporating antibiotic use rates achieved the best prediction performance overall among outcome-only and predictor-enhanced ARIMA, VAR, and LSTM models evaluated (Figure 4, Panel A). Across all model classes, antibiotic-use-enhanced versions yielded lower average RMSEs than their outcome-only counterparts. Model ranking remained consistent across outcomes, with antibiotic-use-enhanced LSTM performing best, followed by antibiotic-use-enhanced VAR and ARIMA models. Among the three models, LSTM consistently showed the greatest benefit from adjustment across all outcomes, with RMSE ratios well below 1 (Figure 4, Panel B).

**Figure 4:**
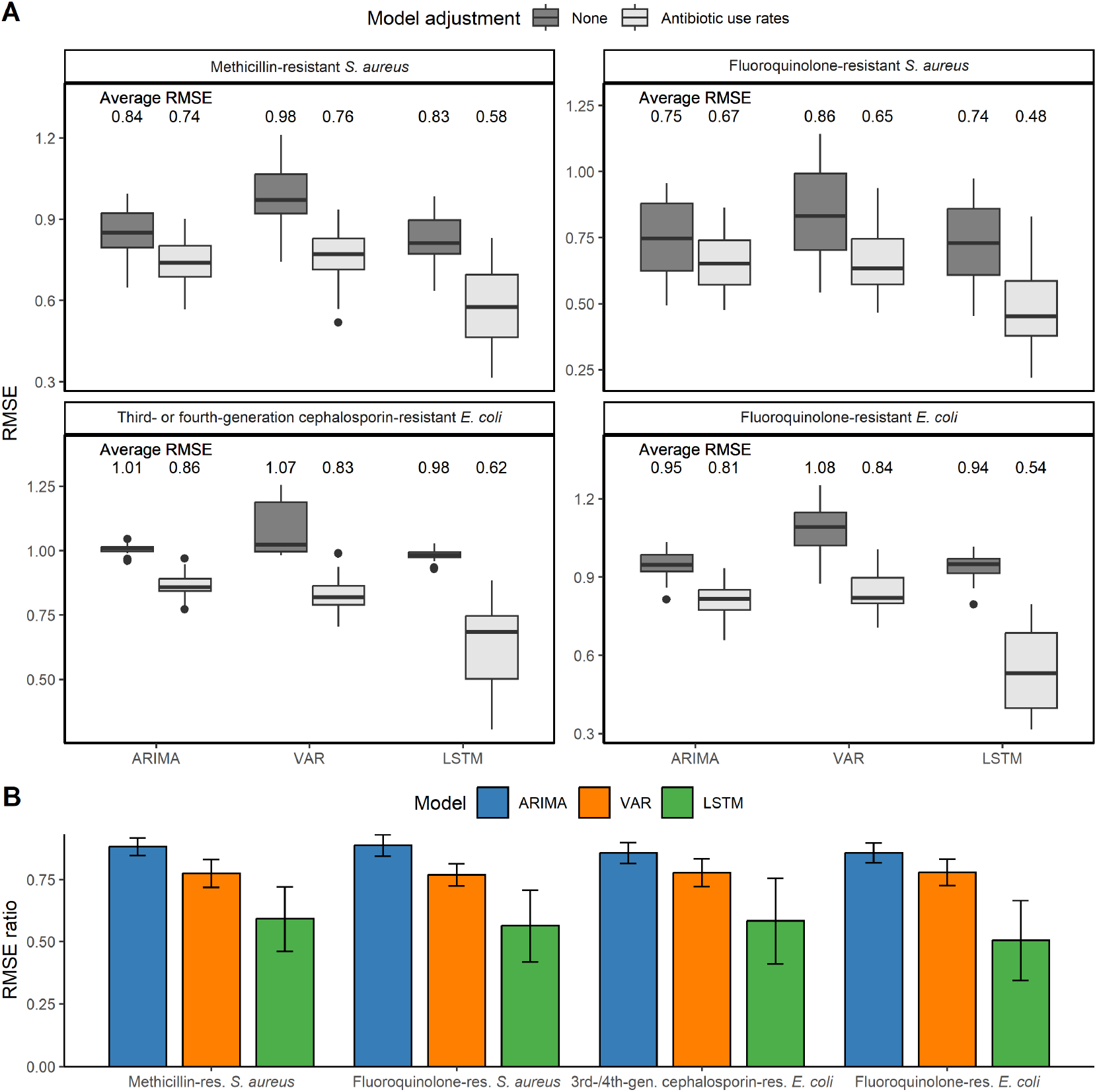
Prediction performance of ARIMA, VAR, and LSTM models across four outcome-specific time series. (A) Prediction performance was measured by root mean squared error (RMSE) on standardized time series. Boxplots show the distribution of RMSE across facility-level time series for each model. Models without adjustment and those incorporating the predictor group of antibiotic use rates are distinguished by the fill color of the boxes (dark vs. light gray). Numerical values above indicate the average RMSE for the outcome-only and predictor-enhanced models. (B) RMSE ratios comparing outcome-only to predictor-enhanced models, stratified by model type. Bars represent the average RMSE ratio across facility-level time series for each outcome, with error bars indicating standard deviations. Lower RMSE ratios suggest greater improvement from adjustment; a value below 1 indicates that adjustment led to lower prediction error.

In zero-out analysis, the removal of Group 1 predictors (i.e., antibiotic use rates) led to the greatest degradation in performance across all outcomes except fluoroquinolone-resistant *S. aureus* (Figure 5, Panel A). In the disaggregated analysis of antibiotic use rates, the predictor with the highest RMSE ratio varied by outcome (Figure 5, Panel B). For both MRSA and fluoroquinolone-resistant *S. aureus*, removal of macrolide use rate resulted in the largest increase in RMSE, Among the *E. coli* outcomes, carbapenem use rate was the most influential for third- or fourth-generation cephalosporin-resistant *E. coli*, while fluoroquinolone use rate had the greatest impact for fluoroquinolone-resistant *E. coli*. These results indicate outcome-specific importance of antibiotic use rates, with certain agents driving prediction performance more strongly depending on the resistance phenotype.

**Figure 5:**
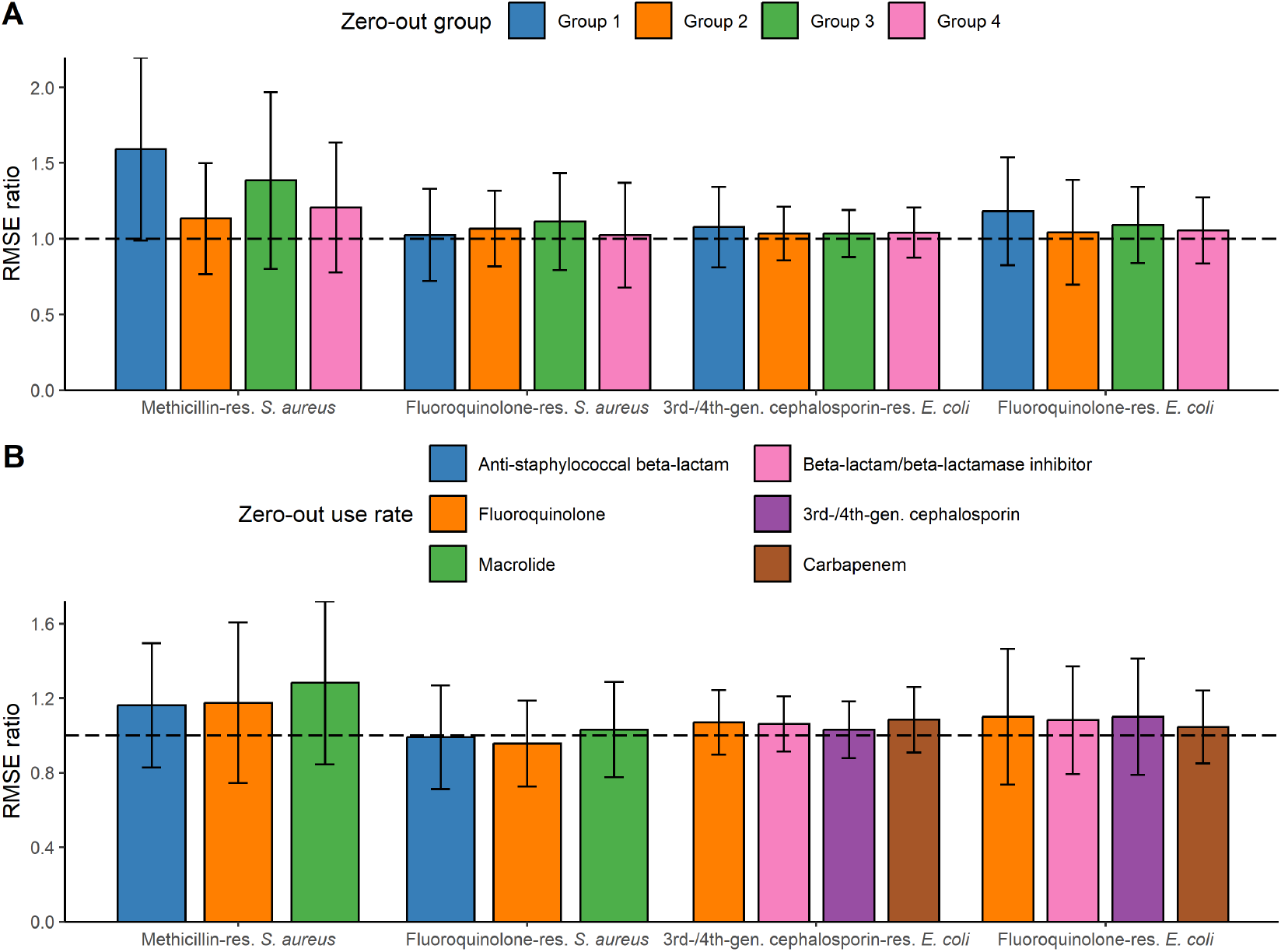
Relative predictive importance of predictors, evaluated by zero-out analysis. (A) RMSE ratios comparing models with specific predictor groups zeroed out to fully predictor-enhanced models. Group 1: antibiotic use rates; Group 2: HOI incidence rates of pathogens; Group 3: COI resistance and susceptibility rates; Group 4: resistant HOI incidence for related antibiotic classes. (B) RMSE ratios when zeroing out individual antibiotic use rate variables. Bars represent the mean RMSE ratio across facility-level time series for each outcome; error bars indicate standard deviations. The horizontal dashed line at 1 represents no change in prediction performance. Values above 1 indicate reduced prediction performance when the predictor group or antibiotic use rate is removed, with larger ratios suggesting greater importance of that predictor or variable for accurate prediction.

## 4 Discussion

This study evaluated and compared three time series forecasting approaches, ARIMA, VAR, and LSTM, for predicting facility-level antibiotic resistance trends in HOIs within the VHA. Our findings demonstrate that LSTM models consistently outperformed classical statistical methods in forecasting accuracy, particularly when incorporating antibiotic use predictors. Moreover, zero-out analysis demonstrated that removal of the antibiotic use group led to the largest degradation in predictive accuracy for all outcomes except fluoroquinolone-resistant *S. aureus*. Macrolide use rates emerged as an important influential predictor for S. aureus resistance phenotypes.

These findings are consistent with prior work showing the superiority of LSTM models over traditional time series approaches in health-related time series, and extend this evidence to the domain of antimicrobial resistance forecasting at the facility level. The consistent ranking of models when enriched with antibiotic use rates suggests that LSTM’s capacity to learn complex temporal dependencies offers a substantial advantage for capturing the nonlinear relationships between prescribing behavior and resistance trends.

Seasonality strength was uniformly weak (medians < 0.3) across outcomes, suggesting that explicit seasonal components may offer limited additional value, whereas trend strength varied markedly, particularly for antibiotic use rates (medians up to 0.80). The predominance of moving average components and limited selection of pure autoregressive structures further underscores the importance of modeling transient shocks and noise rather than long-memory processes in these series. These observations support a parsimonious modeling strategy that prioritizes trend and short-memory smoothing over seasonal decomposition.

The zero-out analysis further confirmed the key role of antibiotic use rates in improving predictive performance, with their removal resulting in a larger increase in RMSE across most outcomes. The importance of specific antibiotic classes varied by pathogen and resistance phenotype, reflecting the nuanced relationship between antibiotic pressure and resistance emergence. For example, macrolide use was more influential for MRSA prediction, while fluoroquinolone use had a larger impact for fluoroquinolone-resistant *E. coli*. These results provide insight into outcome-specific drivers of resistance trends and offer potential targets for stewardship interventions.

Several methodological strengths support the validity of our findings. The zero-out analysis provided insight into the marginal contribution of predictor groups, offering an interpretable measure of variable importance. The use of multiple outcome definitions and diverse predictor groups enabled a comprehensive assessment of model behavior across various resistance scenarios.

Nonetheless, this study has limitations. While the LSTM model was tuned based on prior literature and limited experimentation, more extensive hyperparameter tuning or ensemble approaches may further improve accuracy. Additionally, model performance was assessed using RMSE, which reflects average predictive accuracy but does not quantify uncertainty around forecasts. Incorporating prediction intervals or probabilistic forecasting methods could enhance the interpretability and robustness of future analyses.

Future work should explore the integration of additional facility-level covariates, such as infection control interventions, prescribing practices, or regional epidemiology. Incorporating patient-level data may also improve granularity and support personalized resistance forecasting. Real-time implementation of these models within antimicrobial stewardship programs could facilitate proactive resource allocation and targeted interventions to curb emerging resistance trends.

In summary, our study highlights the potential of LSTM models enhanced with antibiotic use data to provide more accurate forecasts of facility-level resistance trends. These models may serve as valuable tools for data-driven antimicrobial stewardship, enabling proactive responses to emerging resistance threats within healthcare systems like the VHA.

## Data Availability

Individual-level patient data cannot be provided due to Veteran Affairs privacy practices. Aggregated results and analysis code are available upon reasonable request to the authors.

